# Nothing About Us Without Us: A Scoping Review and Priority-Setting Partnership in Type 1 Diabetes and Exercise

**DOI:** 10.1101/19006452

**Authors:** Nika M.D. Klaprat, Nicole Askin, Andrea MacIntosh, Nicole Brunton, Jacqueline L. Hay, Jane E. Yardley, Seth D. Marks, Kathryn M. Sibley, Todd A. Duhamel, Jonathan M. McGavock

## Abstract

**Objectives:** Examine the characteristics of patient engagement (PE) practices in exercise-based randomized trials in type 1 diabetes (T1D), and facilitate T1D stakeholders in determining the top ten list of priorities for exercise research.

**Design:** Two methodological approaches were employed: a scoping review and a modified James Lind Alliance priority-setting partnership.

**Methods:** Published (Medline, Embase, CINAHL, and Central databases) and grey literature (www.clinicaltrials.gov) were searched to identify randomized controlled trials of exercise interventions lasting minimum four weeks and available in English. We extracted information on PE and patient-reported outcomes (PROMs) to identify if patient perspectives had been implemented. Based on results, we set out to determine exercise research priorities as a first step towards a patient-engaged research agenda. An online survey was distributed across Canada to collect research questions from patients, caregivers and healthcare providers. We qualitatively analyzed submitted questions and compiled a long-list that a twelve-person stakeholder steering committee used to identify the top ten priority research questions.

**Results:** Of 9,962 identified sources, 19 published trials and 4 trial registrations fulfilled inclusion criteria. No evidence of PE existed in any included study. Most commonly measured PROMs were frequency of hypoglycemia (n=7) and quality of life (n=4). The priority-setting survey yielded 194 submitted research questions. Steering committee rankings identified 10 priorities focused on lifestyle factors and exercise modifications to maintain short-term glycemic control.

**Conclusion:** Recent exercise-based randomized trials in T1D have not included PE and PROMs. Patient priorities for exercise research have yet to be addressed with adequately designed clinical trials.

## Introduction

Exercise provides numerous health benefits for individuals with type 1 diabetes (T1D)^1^ and is an important component of diabetes self-management^2^. Despite vast health benefits, only one third of people with T1D meet minimum recommendations for regular exercise to achieve health benefits^3^. The unique barriers to exercise for people with T1D^4,5^ are severe, particularly loss of glycemic control and hypoglycemia. With few evidence-based strategies available to overcome these barriers, novel approaches are needed to improve the efficacy of future exercise trials to address patient-relevant concerns.

Including patients in designing and delivering research studies can help address patient-relevant gaps in clinical research^6,7^ such as understanding barriers to uptake of exercise among people with T1D. Patient-oriented research, being “a continuum of research that engages patients as partners, focusses on patient-identified priorities and improves patient outcomes” ^8^, is becoming a priority within clinical trials, but has had little traction in exercise and T1D science^9^. Individuals with T1D have previously been involved in a range of patient engagement^10–12^ (PE) or priority-setting activities^13^ to optimize blood glucose self-management and overall health. Notably, these studies have not centred on exercise research. The current status of PE in setting priorities for and conducting research within exercise science for individuals with T1D remains relatively unknown.

This study addresses these gaps in the literature. First, we conducted a scoping review of exercise training randomized trials for people with T1D to map patient engagement within recent trials. Informed by these results, we then engaged people with T1D, caregivers and healthcare providers in conducting a modified James Lind Alliance (JLA) model^14^ of research prioritization to identify the most important questions about exercise and health.

## STUDY DESIGNS AND RESEARCH METHODS

### Study 1: Scoping Review of Physical Activity/Exercise Randomized Trials and Type 1 Diabetes

We conducted a scoping review of published and grey literature available from the past twenty years to determine in a single narrative analysis: (1) the characteristics of exercise training interventions delivered to people with T1D and (2) the extent patient partners or patient-reported outcomes (PROMs) were involved in study development. The primary question guiding this review was “Is there evidence of patient perspectives being implemented in developing or implementing long-term exercise training trials for individuals with T1D?”

This review was conducted following the five-stage Arksey and O’Malley framework^15^ and formatted in accordance with PRISMA Scoping Review reporting guidelines^16^. A review protocol was not published prior to its conduct. Our team conceptualized PE as per the Canadian Institutes of Health Research definition as being “meaningful and active collaboration in governance, priority-setting, conducting research and knowledge translation” ^8^.

#### Data Sources

Information for this review was collected from published and grey literature. A trained university librarian (NA) developed and implemented search strategies (Appendix A). The published literature search strategy was developed for Medline and adapted to Embase, CINAHL and Central databases respectively. Database searches were conducted on August 22^nd^, 2018, updated on May 16^th^, 2019, and restricted to articles published in the preceding 20 years (January 1998 to May 2019, inclusive). Citations and abstracts for identified publications were uploaded to Rayyan QCRI review management software^17^ for screening.

Additionally, the Clinical Trials online registry (www.clinicaltrials.gov) was searched to identify ongoing trials (grey literature).

Included trial registrations satisfied the same inclusion criteria as published literature according to information provided. Registrations with related publications were added to the published literature analysis; otherwise, detailed aims and protocols were recorded.

#### Inclusion/Exclusion Screening

We included randomized controlled trials of exercise training for individuals with T1D, limiting to interventions lasting four weeks or longer to exclude lab-based acute exercise studies, which are common in exercise science. Interventions providing education to support behaviour change without directly implementing an exercise program were excluded. Full-text sources had to be available in the English language.

Screening of published literature occurred in duplicate. Two reviewers independently screened titles and abstracts from the initial (NK and AM) and updated searches (NK and NB). Conflicts following independent screening were resolved through discussions between reviewers. Full-text versions of potentially eligible articles were searched and uploaded to Rayyan software. Full-text screening was undertaken by both reviewers concurrently. The principal investigator (JM) was consulted throughout screening where disagreement remained after reviewer discussions.

#### Data Extraction

Publications were randomly divided between two co-authors to independently extract data (NK and AM). Where further information was required^18,19^, corresponding authors were contacted electronically. A data extraction form was developed in Microsoft Excel for all published and grey literature data to identify: publication information, participant characteristics, intervention details (frequency, intensity, type, time and intervention duration) and measured outcomes (extracted as per reporting within each study). Reviewers noted evidence of patient engagement if authors declared involvement of people with T1D in research question selection, study design, recruitment, data collection, data analysis and interpretation, or manuscript preparation. Finally, reviewers recorded whether PROMs were measured, as a proxy for reflecting patient-relevant research interests. PROMs included quality of life, diabetes distress, perceived competence, problem areas, self-management behaviours, frequency of glycemic symptoms and several core outcomes identified by the Irish D1 Now Study^12^.

### Study 2: A Priority-Setting Partnership for Research in Type 1 Diabetes and Exercise

Following the scoping review, our team conducted a priority-setting partnership with patients, caregivers and healthcare providers living or working with T1D. We adapted the JLA model of priority-setting, which is supported by the Cochrane Collaboration^20^. This study was approved by the University of Manitoba Health Research Ethics Board (H2018:187) and is reported in accordance with GRIPP2 reporting standards for patient and public involvement in research^21^.

The JLA approach to priority-setting is a multi-stage, mixed-methods research design^14,22^, which we modified to identify priorities for exercise and T1D.

#### Phase 1

We created an online survey using REDCap Surveys server hosted at the University of Manitoba^23,24^ to collect responses to the item, “What questions about physical activity and type 1 diabetes would you like to see answered by research?” Four open-text response boxes were available for respondent submissions. Demographic (age, province of residence and relationship to diabetes) and related patient information (current age, age of diagnosis, gender and ethnic identity) were also collected. The survey was distributed across Canada for six months through partnerships developed with diabetes advocacy organizations (JDRF, Diabetes Canada and Diabetes Action Canada), social media advertising, and posters in diabetes clinics or wellness centres in several urban centres. Concurrently, 12 individuals (eight patients, three caregivers and four healthcare providers) were recruited via maximum variation sampling methods^25^ to participate in a steering committee. This steering committee was responsible for prioritizing submitted questions.

#### Phase 2

Upon survey closure (February 2019), demographic information was extracted directly from the REDCap database. Submitted questions were uploaded to NVivo 12 analysis software. A graduate student trained in qualitative research methods (NK) analyzed submissions by conventional content analysis methodology^26^. Four senior investigators (JMM, TAD, SDM and KMS) were consulted throughout analysis to review results and provide guidance for complicated decisions. A long-list of 38 research questions was developed for phase 3 as per JLA recommendations^14^.

#### Phase 3

Long-listed questions were distributed to the steering committee in a randomized order. Each committee member reviewed the list and ranked their top 10 questions in order of one (most important) to ten (tenth most important). Rankings were returned after the two-week review period by email in word documents encrypted with personalized passwords for each member. Rankings were collated through an inverted points-based system whereby top-ranked questions of each member were denoted ten points, and each successive ranking received one less point. Total points for each question were summed and each question receiving ten or more points (in keeping with the JLA T1D partnership) was short-listed for further prioritization.

#### Phase 4

A one-day in-person workshop for steering committee members was facilitated by the research team (NK, JMM, NB and JLH) to create the final top 10 list of research priorities in exercise and T1D. The goal of the workshop was to reach consensus on priority research questions, defined as every member having at least 80% agreement with the resulting top ten list. The workshop began with ice-breaker activities followed by an independent prioritization activity. Committee members were asked to individually select, in no particular order, their top and bottom three questions from the short-list.

The steering committee was then divided into three groups, each with equal proportions of representative stakeholders. Printed cards for short-listed questions were presented to committee members in categories anonymously reflecting their independent selections (ie. consensus important, less important, mixed ratings or no ratings). Each group ranked all short-listed questions in order of importance. A collated ranking from all groups was presented to members, and the full steering committee was divided into three different groups. These new groups discussed the list and re-ordered printed cards, if necessary.

The final top ten list was presented to the full steering committee. Each member anonymously rated their level of agreement from one to ten. Consensus on the final top ten list was not reached after the first two rounds of group discussions, and a third round was necessary.

### Patient and Public Involvement

As per the IAP2 Spectrum of Public Participation^27^, patients and public stakeholders were consulted to prioritize research questions throughout three of four priority-setting project phases. All research question data were developed and prioritized by patients, caregivers and healthcare providers of individuals with T1D. These research questions will inform our future research agenda, for which we plan to collaborate with stakeholder partners.

## RESULTS

### Study 1: Scoping Review

The published literature search yielded 9,470 citations (Figure 1). Following independent deletion of duplicates and title and abstract screening, 43 citations remained for full-text review. Twenty citations remained after full-text review, two of which were clinical trial registrations and added to the grey literature, leaving 18 published articles. Grey literature searches identified 492 possibly relevant registrations. After eligibility screening, seven fulfilled inclusion criteria; however, four were excluded as relevant articles were already included in the published literature analysis. One registration provided a full-text publication, and was thus added to the published literature. Therefore after screening, 19 published articles and four registered trials were included for analysis.

**FIGURE 1:**
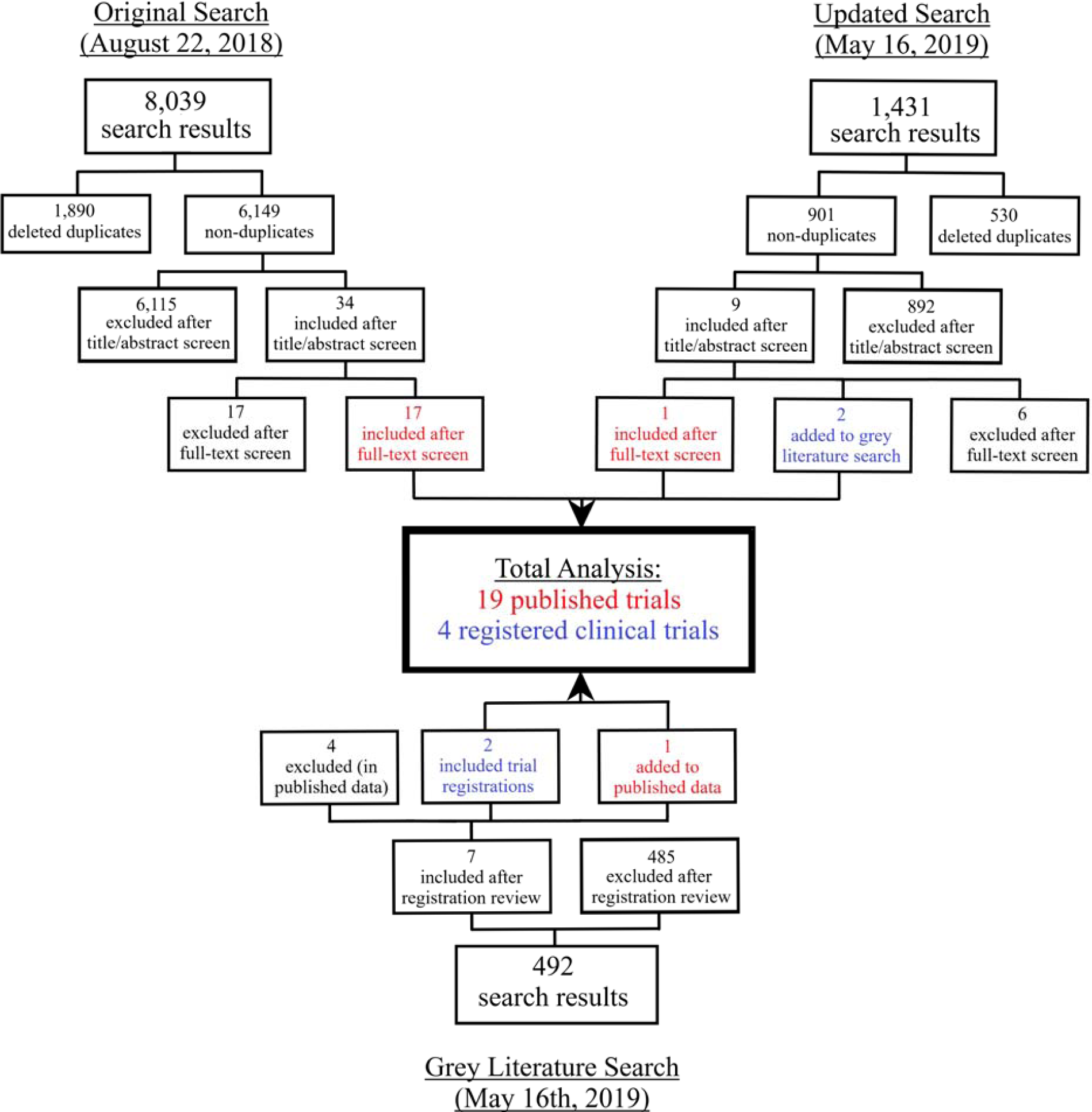
STUDY FLOW DIAGRAM.

#### Published Literature

Data were available for 890 individuals living with T1D (n=18 trials reporting sample sizes). Among studies providing demographic information, 53% of participants were female (n=17 trials), 61% were children or adolescents (n=16 trials) with a mean HbA1c of 8.43% (95% CI: 7.26-9.61%, n=13 trials). Most participants were sedentary or unconditioned (n=12 trials) at enrollment and had lived with T1D for a mean of 5.5 years (median: 5.4; range: 2.9-24.4 years, n=14 studies).

Intervention summaries are provided in Tables 1 and 2. The majority of trials compared aerobic or combined aerobic and resistance training to a non-exercise control group. Interventions were delivered under supervised conditions by a kinesiologist or equivalent, for a median of 60 minutes/session (range: 10-90 minutes), three times/week (range: 1-5 times) for 20 weeks (range: 6 weeks-4 years). Twenty-three outcomes were reported across the 19 trials, of which nearly all focused on physiological factors, with glucose control and predictors of cardiovascular health being most common.

**TABLE 1:**
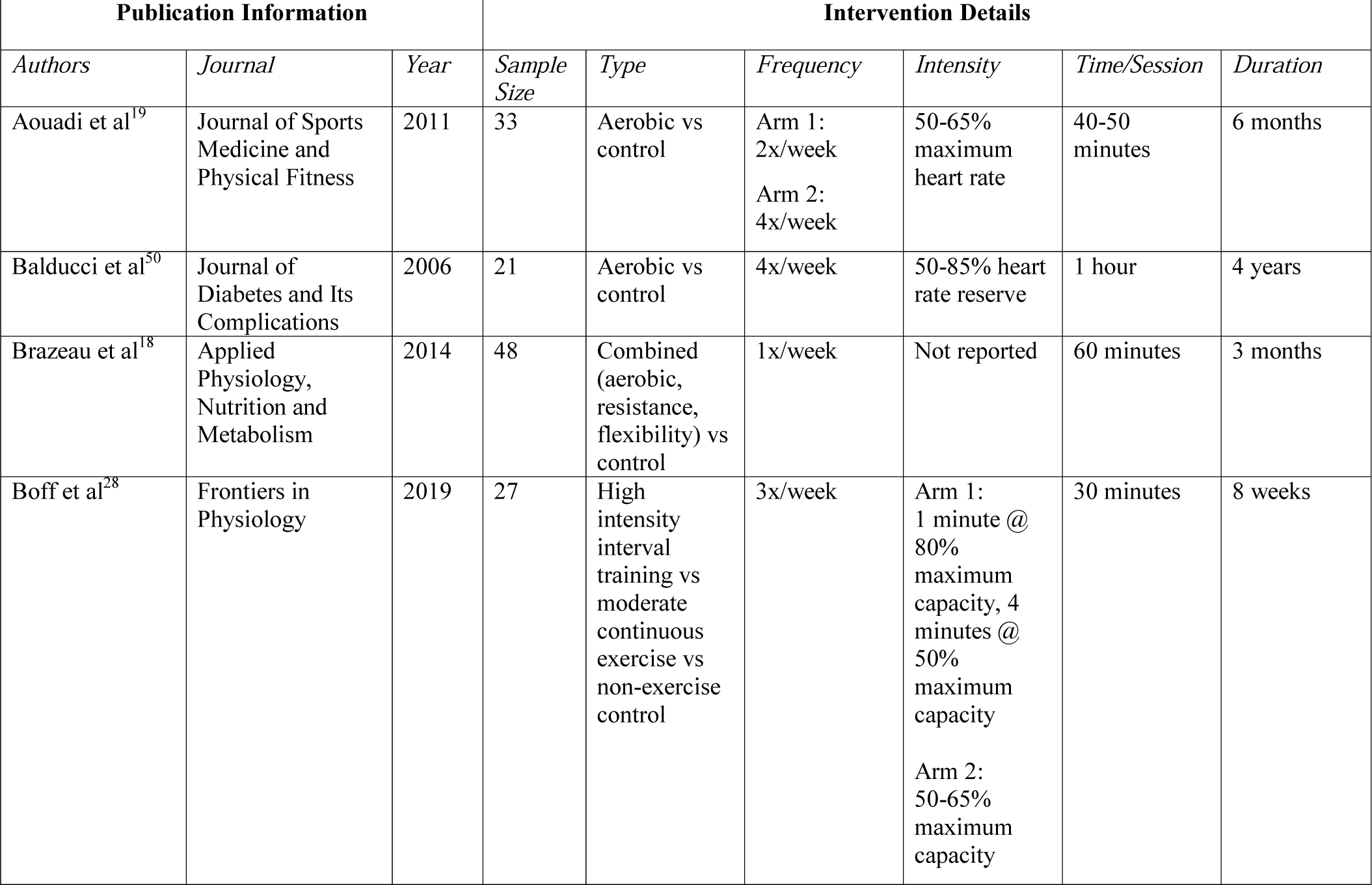
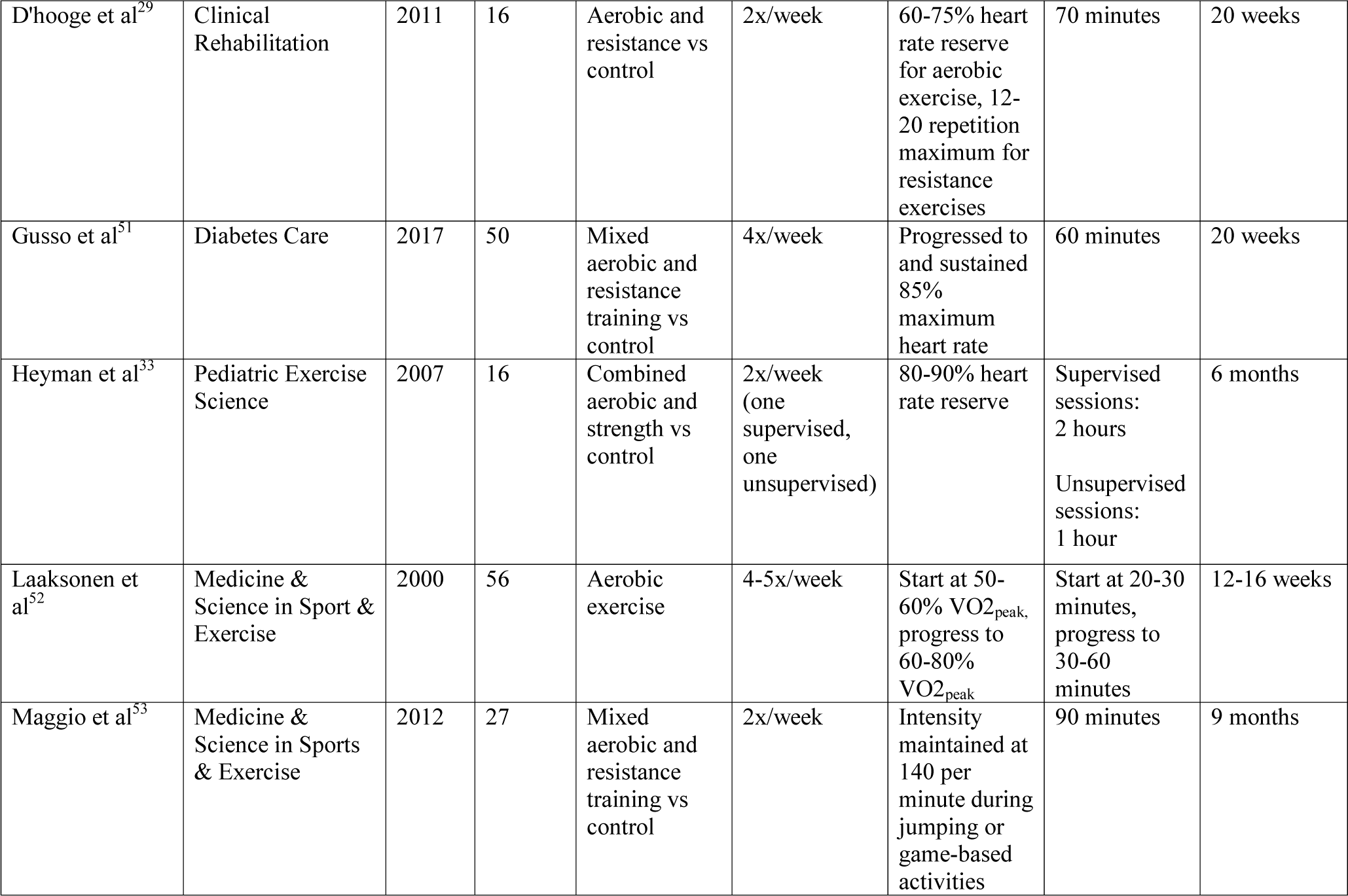
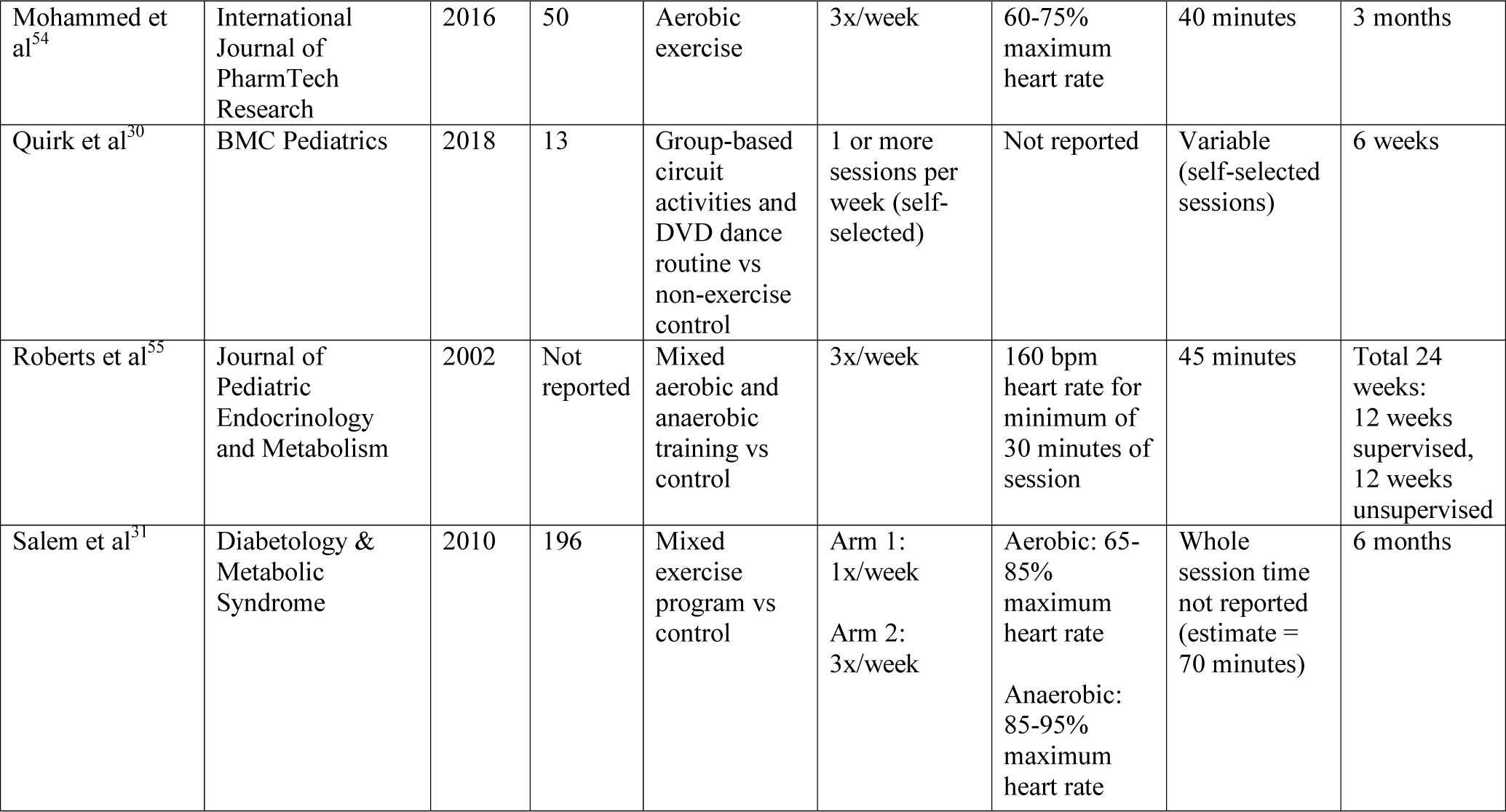
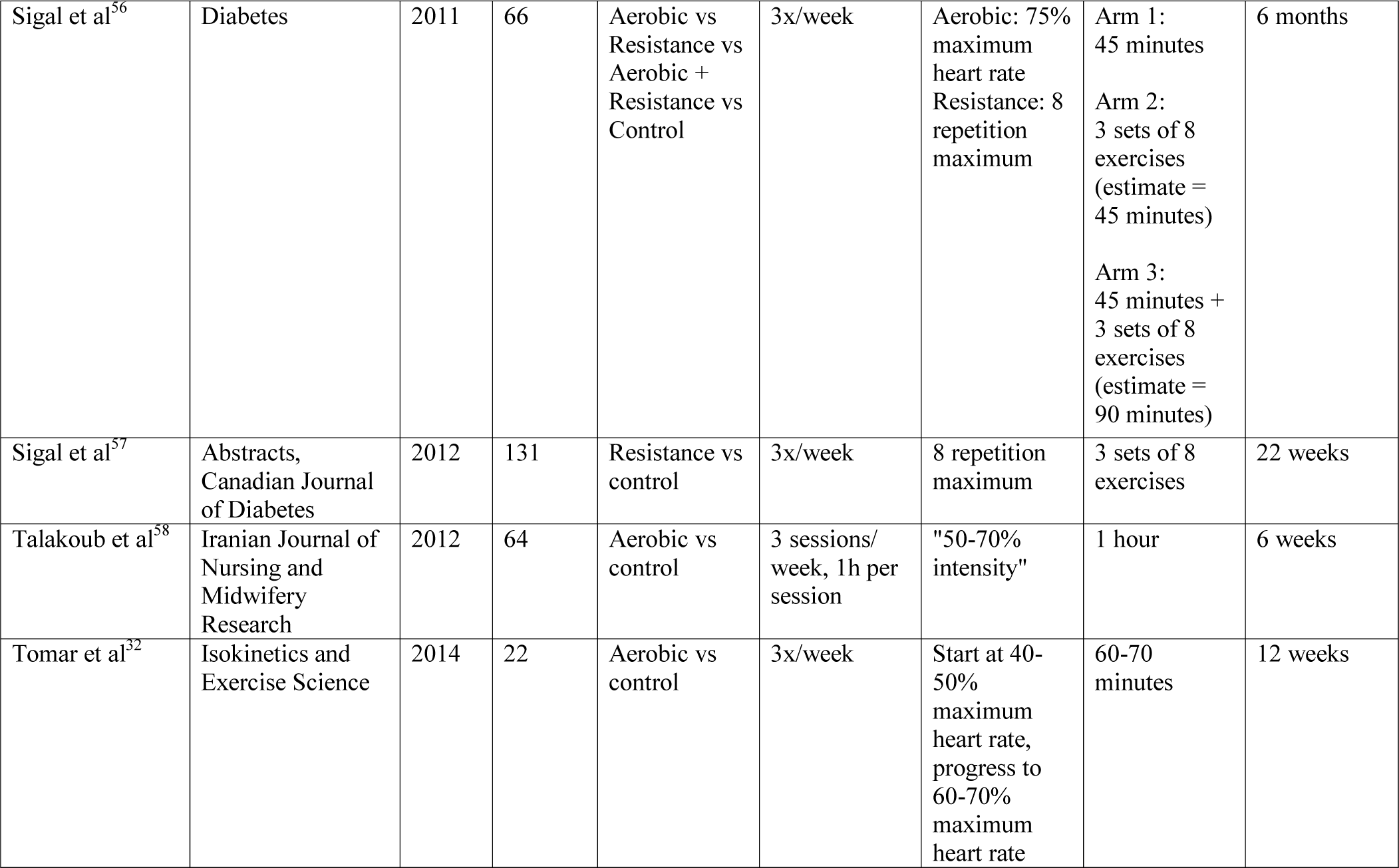
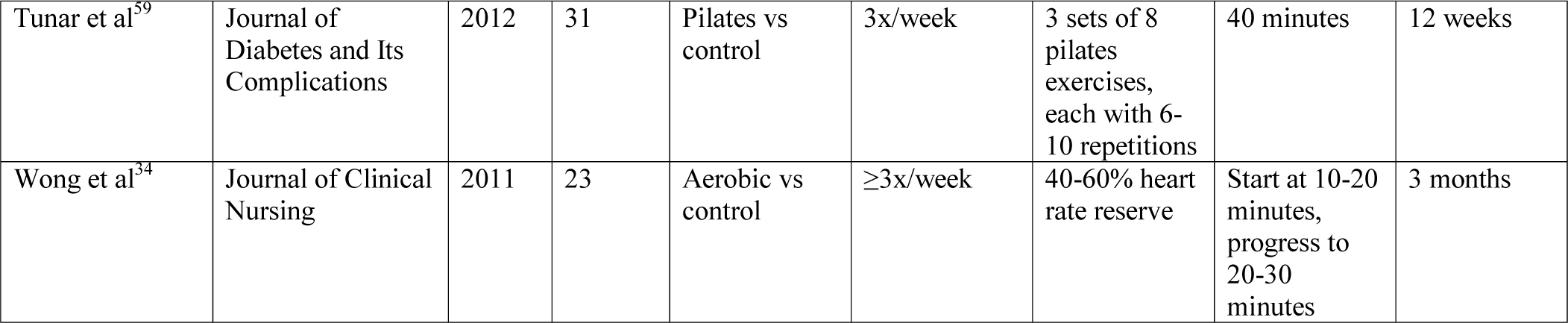
FITT PRINCIPLES OF INCLUDED STUDIES.

**TABLE 2:**
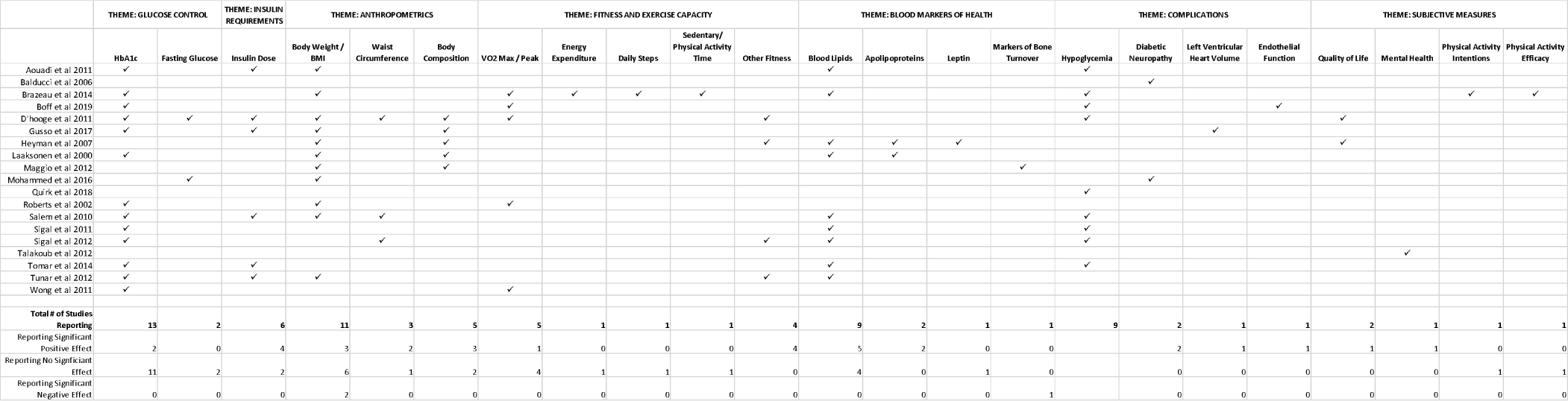
INCLUDED ARTICLES REPORTING OF OUTCOMES.

There was no evidence that any trials conducted to date engaged individuals with lived experience of T1D. Number of hypoglycemic events was the most commonly discussed PROM (n=7 studies)^18,19,28–32^. Additionally, episodes of diabetic ketoacidosis were indirectly observed in two studies^30,32^ (number of adverse events) and quality of life was measured in three studies^29,33,34^ (using different scales).

#### Clinical Trials Registrations

Across the four identified registered clinical trials^35–38^ (Table 3), there are plans to collect data from 187 participants. Three trials are exclusively enrolling youth participants. Planned exercise sessions frequency is a median of 3 times/week (range: 2 times/week – 3x/day, n=3 trials) for 45 minutes/session (range: 3-50 minutes, n=3 trials) lasting 15.1 weeks (range: 12-32 weeks, n=4 trials). One trial^37^ has reached target enrollment and anticipates publishing by the end of 2019. No trial registration described partnerships with stakeholders in developing or implementing the study. Quality of life is the only PROM explicitly disclosed in one trial^38^.

**TABLE 3:** INCLUDED REGISTERED CLINICAL TRIALS.

| Registration Information |  |  |  | Intervention Details |  |  |  | Measured Outcomes |  |
| --- | --- | --- | --- | --- | --- | --- | --- | --- | --- |
| Principal Investigator | Registration # | Status | Last Update | Sample Size | Type | Duration | Frequency | Primary | Secondary |
| Pierre Fontaine | clinicaltrials.gov:<br>NCT03528226 | Not yet recruiting | 15/Oct/18 | 34 | Aerobic | 4 months | 2x supervised,<br>1x un-supervised<br>per week | 1. Flow mediated dilation | 1. Vascular responses<br>2. VO<sub>2</sub><sub>max</sub><br>3. Blood nitric oxide and neurotrophic factors<br>4. Body composition |
| Dominique Daurman | clinicaltrials.gov:<br>NCT03199638 | Unknown | 27/Jun/17 | 24 | Exercise snacks | 3 months | 21x/week<br>(3x/day) | 1. HbA1c<br>2. MAGE<br>3. % time blood glucose in/above/below target range<br>4. Insulin sensitivity<br>5. Insulin dose | N/A |
| Ramon Baron | WHO Registry:<br>ISRCTN12066515 | Complete | 1/Jul/19 | 29 | Resistance | 32 weeks | 2x/week | 1. HbA1c<br>2. Physical fitness (6 minute walk test and strength tests) | 1. Adiponectin levels<br>2. Intima media thickness of left carotid artery |
| Palak Kishorbhai Purohit | WHO Registry:<br>CTRI/2018/03/012270 | Not yet recruiting | 5/Mar/18 | 100 | Yoga | 12 weeks | Not specified | 1. Signs/symptoms of juvenile diabetes | 1. Prevention of complications<br>2. General health status<br>3. Insulin dose<br>4. Quality of life |

### Study #2 - Priority-Setting Exercise

The online survey was available and advertised to the public between July 2018 and January 2019. We collected responses from 115 individuals across nine Canadian provinces. Respondents were a mean age of 40.9 (±15.1) years, and the majority (73.9%) identified as a patient with T1D. The remainder identified as caregivers (15.7%), friends (7.0%) or healthcare providers (12.2%), with some respondents identifying as more than one category (8.7%). More females completed the survey (63.4%) than males, and no participants identified as a non-binary gender. Most patients identified as being of Canadian ethnic origin (74%), with the next largest samples being European Canadian (15%), Métis (4%) and Caribbean Canadian (3%).

Of the 115 respondents, 100 submitted at least one research question, producing a total of 194 submissions. After qualitative analysis (Figure 2), 38 research questions were included in the phase 3 long-list and distributed to steering committee members for review. We received 100% of our steering committee rankings between February 21^st^ and March 8^th^, 2019. Twenty-four questions were short-listed after receiving ten or more points in collated rankings.

**FIGURE 2:**
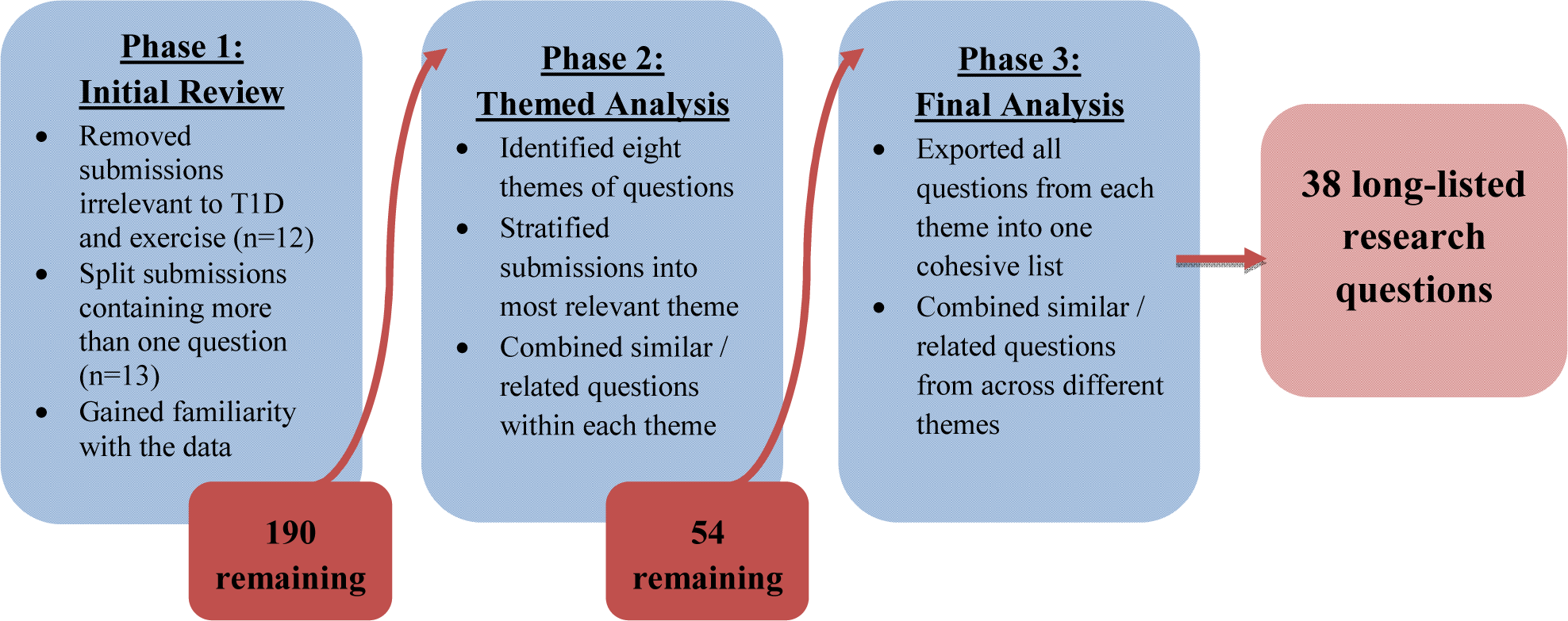
QUALITATIVE ANALYSIS PROCESS.

Eleven of twelve steering committee members attended the Phase 4 workshop on April 6^th^, 2019. The workshop lasted approximately six hours. Three rounds of small group discussions occurred. Following the second round, three members did not sufficiently agree with the aggregated list. The third round resulted in slightly lower agreement for two of those members. Therefore, we conducted a post-workshop analysis using small group rankings from the third round to supplement the aggregated list from the second round. This analysis resulted in the final top ten list of research priorities for T1D and exercise (Table 4).

**TABLE 4:** RESEARCH PRIORITIES FOR EXERCISE AND TYPE 1 DIABETES.

| Position # | Research Question |
| --- | --- |
| 1 | What explains the variation in responses that the same person can experience doing the same exercise between different days? |
| 2 | Which is the best for maintaining glycemic stability and glucose tolerance: aerobic training, strength training, or a combination of both? If a combination, does the order matter? |
| 3 | What modes of exercise (ie activity types, such as walking, cycling, weightlifting, rock climbing etc) produce the best health benefits while maintaining tight glycemic control? |
| 4 | What dietary plans can safely and effectively be followed for an active lifestyle in type 1 diabetes without compromising pre- and post-exercise glycemic control? |
| 5 | What is the optimal time of day and exercise prescription (example: how often, what type, how intense) in order to maintain ideal glycemic control and insulin sensitivity? |
| 6 | What is the best method of preventing post-exercise hypo- or hyperglycemia? |
| 7 | Will certain glycemic ranges before starting exercise consistently result in hypo- or hyperglycemia? |
| 8 | What effect can various levels of hydration have on blood sugar levels during and after exercise? |
| 9 | How does hypo- or hyperglycemia affect muscle growth and strength training progress, or vice versa? |
| 10 | What is the effect of climate/temperature on blood sugar control during exercise and what causes this effect? |

## DISCUSSION

This study examined the characteristics of randomized trials and patient priorities for exercise science research for people with T1D. In our scoping review, we determined that patient engagement methods and PROMs have not been historically used to inform exercise-based interventions. Guided by these results, we facilitated a priority-setting project with T1D stakeholders to identify priority research questions pertaining to exercise and health. We identified that patients and caregivers are interested in modalities and strategies to exercise safely and maintain glucose control. Collectively, these findings provide a novel patient-centred rationale for designing future randomized trials of exercise interventions for people with T1D.

### Previous Literature

This is the first scoping review of exercise randomized trials for individuals with T1D designed to determine if patient engagement exists in exercise and T1D literature. This topic was not addressed in recent systematic reviews of exercise training and health outcomes in people with T1D^39–41^. We found that exercise randomized trials published or being delivered for individuals with T1D did not focus on stakeholder engagement. This gap is not exclusive to trials of T1D and exercise. A scoping review of priority-setting practices in all health research found only 27studies engaged patients in identifying research topics, and 12 in identifying specific research questions^42^. Many studies simply inferred stakeholder priorities from qualitative data. Additionally, most trials engaging stakeholders do not integrate multiple stakeholders’ perspectives (ie. patients, clinicians, caregivers etc) in the prioritization process. This is an important consideration when engaging stakeholders in research, as stakeholders with different experiences of a health condition may have different priorities for research topics or outcomes^43^. Engaging T1D stakeholders is a significant gap in exercise science literature and should be considered within future randomized trials.

Patient engagement and priority-setting projects identifying important research topics from stakeholders’ perspectives are becoming more common within clinical research^42,44,45^. This project revealed that stakeholders are largely concerned with short-term outcomes, strategies to prevent hypoglycemia and stabilize short-term glucose control. This contrasts the JLA T1D treatments project^13^, where prioritized questions focused on long-term outcomes including adverse effects of various insulin analogues or potential cognitive impacts of living with T1D. This difference may indicate some uncertainty felt by stakeholders regarding the safety of exercise given their individual situation. Fear of short-term health complications is a common barrier to regular exercise among people with T1D^5^. This fear itself has a range of health implications including reduced physical activity^46^, increased glycemic variability^46^, poorer sleep patterns^47^ and reduced quality of life^47^. Although guidelines and consensus statements about prevention of post-exercise hypoglycemia exist^48,49^, the literature on which these recommendations are based has limitations. As this is the first investigation into patient priorities in T1D and exercise, previous research may not have been intentionally directed towards established patient-identified questions. Future randomized trials should focus on stakeholder priorities to provide optimally relevant recommendations to individuals with T1D.

### Strengths and Weaknesses

This study was strengthened by using two complimentary methods to identify gaps in exercise science for individuals with T1D. Our priority-setting project followed a modified JLA approach to priority-setting. We believe the recognition and support for this model^20^ is a strength for our study. We were also supported by a highly engaged steering committee (100% phase 3 participation rate). Despite these strengths, several limitations should be addressed. The scoping review was limited to trials published recently and in the English language, therefore we may have missed some trials. Additionally, we were not able to achieve consensus in person at the workshop. Although the JLA Guidebook^14^ mentions a majority vote can be obtained in cases where consensus is not achieved, this process had to be conducted in a post-workshop analysis since several members had other commitments.

### Impact of Patient Engagement

A quantitative evaluation of patient engagement was not conducted. Consulting end users as participants was integral to this study. The developed list of research questions was based on submissions collected directly from and prioritized by end users of research. Without input from patients and caregivers, prioritized research questions would likely have been very different.

## CONCLUSION

We have outlined the current status of patient engagement in exercise research for individuals with T1D and engaged stakeholders in developing a list of priorities in T1D and exercise research. This list of priorities will be used to guide our future research agenda, and we recognize the need to continue working with stakeholders in designing future research. It will also be critical to re-evaluate priorities as new information becomes available.

**Summary Box**

- We found no evidence of patient engagement or involvement in exercise science research for patients living with type 1 diabetes over the past twenty years
- We worked with a Canadian sample of patients, caregivers and healthcare providers living or working with type 1 diabetes to develop a top ten list of priorities for future exercise science research
- Current researchers interested in type 1 diabetes and exercise now have a framework of priorities set by stakeholders, which can be used to guide future research programs and result in more clinically relevant exercise science findings

## Data Availability

The de-identified datasets used and/or analyzed during the current study are available from the corresponding author on reasonable request.

## Statements

### Contributorship statement

NK, JM, NA, AM and NB all contributed to the conduct of the scoping review. NK, JM, TD, KMS, SM, JY, JLH and NB contributed to the priority-setting project, in varying capacities including project planning, data collection and recruitment and data analysis. All authors contributed to writing or editing this manuscript.

### Competing interests and funding statement

This study was supported by funding from the Canadian Institutes of Health Research Strategy for Patient-Oriented Research (SCA-145101) and an Applied Public Health Chair in Resilience and Obesity in Youth held by Dr. McGavock. JLH was supported by a CIHR Vanier Scholarship. KMS holds a Canada Research Chair in Integrated Knowledge Translation in Rehabilitation Sciences. JEY received personal fees and/or non-financial support from Dexcom Canada, Abbott Nutrition Canada and LifeScan Canada, outside the submitted work. NK was supported by a University of Manitoba Graduate Fellowship award. The authors do not declare any competing interests in the conduct of these studies.

### Ethical approval statement

Ethics approval was not required for the scoping review. The priority-setting study was approved by the University of Manitoba Health Research Ethics Board (H2018:187).

## Notes

### Competing Interest Statement

Dr. Yardley reports personal fees and non-financial support from Dexcom Canada, non-financial support from Abbott Nutrition Canada, non-financial support from LifeScan Canada, outside the submitted work. Dr. Hay reports grants from Canadian Institute of Health Research Vanier Scholarship, during the conduct of the study.

### Author Declarations

All relevant ethical guidelines have been followed and any necessary IRB and/or ethics committee approvals have been obtained.

Any clinical trials involved have been registered with an ICMJE-approved registry such as ClinicalTrials.gov and the trial ID is included in the manuscript.

## References

1. Chimen M, Kennedy A, Nirantharakumar K, Pang TT, Andrews R, Narendran P. What are the health benefits of physical activity in type 1 diabetes mellitus? A literature review. Diabetologia. 2012;55(3):542–551. doi:10.1007/s00125-011-2403-2

2. Joslin E. Diabetes. In: Fishbein M, ed. Modern Home Medical Advice: Your Health and How to Preserve It. Garden City, New York: Country Life Press; 1935:563–606.

3. McCarthy MM, Funk M, Grey M. Cardiovascular health in adults with type 1 diabetes. Prev Med (Baltim). 2016;91:138–143. doi:10.1016/j.ypmed.2016.08.019

4. Lascar N, Kennedy A, Hancock B, et al. Attitudes and barriers to exercise in adults with type 1 diabetes (T1DM) and how best to address them: A qualitative study. PLoS One. 2014;9(9). doi:10.1371/journal.pone.0108019

5. Brazeau A-S, Strychar I, Rabasa-Lhoret R, Mirescu H. Barriers to Physical Activity Among Patients With Type 1 Diabetes. Diabetes Care. 2008;31(11):2108–2109. doi:10.2337/dc08-0720.

6. Concannon TW, Fuster M, Saunders T, et al. A Systematic Review of Stakeholder Engagement in Comparative Effectiveness and Patient-Centered Outcomes Research. J Gen Intern Med. 2014;29(12):1692–1701. doi:10.1007/s11606-014-2878-x

7. Brett J, Staniszewska S, Mockford C, et al. A Systematic Review of the Impact of Patient and Public Involvement on Service Users, Researchers and Communities. Patient. 2014;7(4):387–395. doi:10.1007/s40271-014-0065-0

8. Canadian Institutes of Health Research. Strategy for Patient-Oriented Research: Patient Engagement Framework.; 2019. http://www.cihr-irsc.gc.ca/e/documents/spor_framework-en.pdf.

9. Klaprat N, MacIntosh A, McGavock JM. Gaps in Knowledge and the Need for Patient-Partners in Research Related to Physical Activity and Type 1 Diabetes: A Narrative Review. Front Endocrinol (Lausanne). 2019;10(February):1–12. doi:10.3389/fendo.2019.00042

10. Price KJ, Knowles JA, Fox M, et al. Effectiveness of the Kids in Control of Food (KICk-OFF) structured education course for 11-16 year olds with Type 1 diabetes. Diabet Med. 2016;33(2):192–203. doi:10.1111/dme.12881

11. Cafazzo JA, Casselman M, Hamming N, Katzman DK, Palmert MR. Design of an mHealth app for the self-management of adolescent type 1 diabetes: A pilot study. J Med Internet Res. 2012;14(3). doi:10.2196/jmir.2058

12. Byrne M, O’Connell A, Egan AM, et al. A core outcomes set for clinical trials of interventions for young adults with type 1 diabetes: An international, multi-perspective Delphi consensus study. Trials. 2017;18(1):1–8. doi:10.1186/s13063-017-2364-y

13. Gadsby R, Snow R, Daly AC, et al. Setting research priorities for Type 1 diabetes. Diabet Med. 2012;29(10):1321–1326. doi:10.1111/j.1464-5491.2012.03755.x

14. National Institute for Health Research. The James Lind Alliance Guidebook.; 2018. www.jla.nihr.ac.uk.

15. Arksey H, O’Malley L. Scoping studies: towards a methodological framework. Int J Soc Res Methodol. 2005;8(1):19–32. doi:10.1080/1364557032000119616

16. Tricco AC, Lillie E, Zarin W, et al. PRISMA extension for scoping reviews (PRISMA-ScR): Checklist and explanation. Ann Intern Med. 2018;169(7):467–473. doi:10.7326/M18-0850

17. Ouzzani M, Hammady H, Fedorowicz Z, Elmagarmid A. Rayyan — a web and mobile app for systematic reviews. Syst Rev. 2016;5(210). doi:10.1186/s13643-016-0384-4

18. Brazeau A-S, Gingras V, Leroux C, et al. A pilot program for physical exercise promotion in adults with type 1 diabetes: the PEP-1 program. Appl Physiol Nutr Metab. 2014;39(4):465–471. doi:10.1139/apnm-2013-0287

19. Aouadi R, Khlifa R, Aouidet A, et al. Aerobic training programs and glycemic control in diabetic children in relation to exercise frequency. J Sports Med Phys Fitness. 2011;51(1-2):1–8.

20. Khan N, Bacon SL, Khan S, et al. Hypertension management research priorities from patients, caregivers, and healthcare providers: A report from the Hypertension Canada Priority Setting Partnership Group. J Clin Hypertens. 2017;19(11):1063–1069. doi:10.1111/jch.13091

21. Staniszewska S, Brett J, Simera I, et al. GRIPP2 reporting checklists: Tools to improve reporting of patient and public involvement in research. BMJ. 2017;358(j3453):1–7. doi:10.1136/bmj.j3453

22. Lophatananon A, Tyndale-Biscoe S, Malcolm E, et al. The James Lind Alliance Approach To Priority Setting for Prostate Cancer Research: an Integrative Methodology Based on Patient and Clinician Participation. BJU Int. 2011;108(7):1040–1043. doi:10.1111/j.1464-410x.2011.10609.x

23. Harris PA, Taylor R, Thielke R, Payne J, Gonzalez N, Conde JG. Research electronic data capture (REDCap)-A metadata-driven methodology and workflow process for providing translational research informatics support. J Biomed Inform. 2009;42(2):377–381. doi:10.1016/j.jbi.2008.08.010

24. Harris PA, Taylor R, Minor BL, et al. The REDCap consortium: Building an international community of software platform partners. J Biomed Inform. 2019;95(April):103208. doi:10.1016/j.jbi.2019.103208

25. Markula P, Silk M. Practice and the Politics of Interpretation: Interviewing. In: Qualitative Research for Physical Culture. New York, NY: Palgrave MacMillan; 2011:81–111.

26. Hsieh H-F, Shannon SE. Three Approaches to Qualitative Content Analysis. Qual Health Res. 2005;15(9):1277–1288. doi:10.1177/1049732305276687

27. International Association for Public Participation. IAP2 Spectrum of Public Participation.; 2018. https://cdn.ymaws.com/www.iap2.org/resource/resmgr/pillars/Spectrum_8.5X11_Print.pdf.

28. Boff W, da Silva AM, Farinha JB, et al. Superior Effects of High-Intensity Interval vs. Moderate-Intensity Continuous Training on Endothelial Function and Cardiorespiratory Fitness in Patients With Type 1 Diabetes: A Randomized Controlled Trial. Front Physiol. 2019;10(April):1-10. doi:10.3389/fphys.2019.00450

29. D’hooge R, Hellinckx T, Van Laethem C, et al. Influence of combined aerobic and resistance training on metabolic control, cardiovascular fitness and quality of life in adolescents with type 1 diabetes: A randomized controlled trial. Clin Rehabil. 2011;25:349–359. doi:10.1177/0269215510386254

30. Quirk H, Glazebrook C, Blake H. A physical activity intervention for children with type 1 diabetes-steps to active kids with diabetes (STAK-D): A feasibility study. BMC Pediatr. 2018;18(1):1–12. doi:10.1186/s12887-018-1036-8

31. Salem MA, AboElAsrar MA, Albarbary NS, ElHilaly RA, Refaat, Yara M. Is exercise a therapeutic tool for improvement of cardiovascular risk factors in adolescents with type 1 diabetes mellitus? A randomised controlled trial. Diabetol Metab Syndr. 2010;2(47):1–10. http://ovidsp.ovid.com/ovidweb.cgi?T=JS&PAGE=reference&D=emed9&NEWS=N&AN=2010437726.

32. Tomar R, Hamdan M, Al-Qahtani MH. Effect of low to moderate intensity walking and cycling on glycaemic and metabolic control in type 1 diabetes mellitus adolescent males: A randomized controlled trial. Isokinet Exerc Sci. 2014;22(3):237–243. doi:10.3233/IES-140544

33. Heyman E, Toutain C, Delamarche P, et al. Exercise Training and Cardiovascular Risk Factors in Type 1 Diabetic Adolescent Girls. Pediatr Exerc Sci. 2007;19:408–419. doi:10.1123/pes.19.4.408

34. Wong CH, Chiang YC, Wai JPM, et al. Effects of a home-based aerobic exercise programme in children with type 1 diabetes mellitus. J Clin Nurs. 2011;20(5-6):681–691. doi:10.1111/j.1365-2702.2010.03533.x

35. Daurman D. Exercise Snacks and Glutamine to Improve Glucose Control in Adolescents With Type 1 Diabetes. NCT03199638. https://clinicaltrials.gov/ct2/show/NCT03199638%0D. Published 2017.

36. Fontaine P. Exercise Training and Endothelial Function in Type 1 Diabetes (EVaDia). NCT03528226. https://clinicaltrials.gov/ct2/show/NCT03528226. Published 2018.

37. Baron R. Regular resistance training in children with type 1 diabetes improves glycaemic control and physical fitness. ISRCTN12066515. http://www.isrctn.com/ISRCTN12066515. Published 2019.

38. Purohit PK. Management of Jataja Prameha (JUvenile Diabetes) with Modified Nisha Amalaki Yoga. ctri/2018/03/012270. http://apps.who.int/trialsearch/Trial2.aspx?TrialID=CTRI/2018/03/012270. Published 2018.

39. Aljawarneh YM, Wardell DW, Wood GL, Rozmus CL. A Systematic Review of Physical Activity and Exercise on Physiological and Biochemical Outcomes in Children and Adolescents With Type 1 Diabetes. J Nurs Scholarsh. 2019;51(3):337–345. doi:10.1111/jnu.12472

40. Yardley JE, Hay J, Abou-Setta AM, Marks SD, McGavock J. A systematic review and meta-analysis of exercise interventions in adults with type 1 diabetes. Diabetes Res Clin Pract. 2014;106(3). doi:10.1016/j.diabres.2014.09.038

41. Quirk H, Blake H, Tennyson R, Randell TL, Glazebrook C. Physical activity interventions in children and young people with Type 1 diabetes mellitus: A systematic review with meta-analysis. Diabet Med. 2014;31(10). doi:10.1111/dme.12531

42. Stewart RJ, Caird J, Oliver K, Oliver S. Patients’ and clinicians’ research priorities. Heal Expect. 2010;14:439–448. doi:10.1111/j.1369-7625.2010.00648.x

43. Boote J, Baird W, Beecroft C. Public involvement at the design stage of primary health research: A narrative review of case examples. Health Policy (New York). 2010;95:10–23. doi:10.1016/j.healthpol.2009.11.007

44. Manafò E, Petermann L, Vandall-Walker V, Mason-Lai P. Patient and public engagement in priority setting: A systematic rapid review of the literature. PLoS One. 2018;13(3):1–18. doi:10.1371/journal.pone.0193579

45. Barello S, Graffigna G, Vegni E. Patient Engagement as an Emerging Challenge for Healthcare Services: Mapping the Literature. Nurs Res Pract. 2012;2012:1–7. doi:10.1155/2012/905934

46. Martyn-Nemeth P, Quinn L, Penckofer S, Park C, Hofer V, Burke L. Fear of hypoglycemia: Influence on glycemic variability and self-management behavior in young adults with type 1 diabetes. J Diabetes Complications. 2017;31(4):735–741. doi:10.1016/j.jdiacomp.2016.12.015

47. Martyn-Nemeth P, Schwarz Farabi S, Mihailescu D, Nemeth J, Quinn L. Fear of hypoglycemia in adults with type 1 diabetes: Impact of therapeutic advances and strategies for prevention - A review. J Diabetes Complications. 2016;30(1):167–177. doi:10.1016/j.jdiacomp.2015.09.003

48. Riddell MC, Gallen IW, Smart CE, et al. Exercise management in type 1 diabetes: a consensus statement. Lancet Diabetes Endocrinol. 2017;5(5):377–390. doi:10.1016/S2213-8587(17)30014-1

49. Yardley JE, Sigal RJ. Exercise strategies for hypoglycemia prevention in individuals with type 1 diabetes. Diabetes Spectr. 2015;28(1):32–38. doi:10.2337/diaspect.28.1.32

50. Balducci S, Iacobellis G, Parisi L, et al. Exercise training can modify the natural history of diabetic peripheral neuropathy. J Diabetes Complications. 2006;20(4):216–223. doi:10.1016/j.jdiacomp.2005.07.005

51. Gusso S, Pinto T, Baldi JC, et al. Exercise training improves but does not normalize left ventricular systolic and diastolic function in adolescents with type 1 diabetes. Diabetes Care. 2017;40(9):1264–1272. doi:10.2337/dc16-2347

52. Laaksonen DE, Atalay M, Niskanen LK, et al. Aerobic exercise and the lipid profile in type 1 diabetic men: A randomized controlled trial. Med Sci Sports Exerc. 2000;32(9):1541–1548. doi:10.1097/00005768-200009000-00003

53. Maggio ABR, Rizzoli RR, Marchand LM, Ferrari S, Beghetti M, Farpour-Lambert NJ. Physical activity increases bone mineral density in children with type 1 diabetes. Med Sci Sports Exerc. 2012;44(7):1206–1211. doi:10.1249/MSS.0b013e3182496a25

54. Mohammed MA, Rahmy AF, Mohamen GS, Kaddah AF. Effect of exercise training on cardiovascular responses in diabetic autonomic neuropathy. Int J PharmTech Res. 2016;9(5):110–118.

55. Roberts L, Jones TW, Fournier PA. Exercise Training and Glycemic Control in Adolescents with Poorly Controlled Type 1 Diabetes Mellitus. J Pediatr Endocrinol Metab. 2002;15(5):621–627.

56. Sigal RJ, Kenny G, Goldfield G, et al. Neither Aerobic Exercise Nor Resistance Exercise Improves Glycemic Control in Type 1 Diabetes: A Randomized Controlled Trial. Diabetes. 2011;60(0):A38.

57. Sigal RJ, Kenny GP, Perkins BA, et al. Resistance Exercise in Already-active Type 1 Diabetic Individuals: The READI Trial. Can J Diabetes. 2012;36(5):S15. doi:10.1016/j.jcjd.2012.07.065

58. Talakoub S, Gorbani S, Hasanpour M, Zolaktaf V, Amini M. Impact of exercise on affective responses in female adolescents with type I diabetes. Iran J Nurs Midwifery Res. 2012;17(6):434–439.

59. Tunar M, Ozen S, Goksen D, Asar G, Bediz CS, Darcan S. The effects of Pilates on metabolic control and physical performance in adolescents with type 1 diabetes mellitus. J Diabetes Complications. 2012;26(4):348–351. doi:10.1016/j.jdiacomp.2012.04.006

